# Investigating Nutritional Interventions for Managing Irritable-Bowel Syndrome: A Bibliometric Analysis

**DOI:** 10.1101/2024.11.29.24318034

**Authors:** Madho Mal, Faraz Arshad, Sadaf Iftikhar, Nayanika Tummala, Saad Ahmed Waqas, Usama Hussain Kamal, Rizwana Noor, Muhammad Khalid Afridi, Abraish Ali

## Abstract

**Background:** The bibliometric analysis provides a structured approach for providers and researchers to access clinically relevant information to improve patient care.

**Methods:** We used SCOPUS as our database and made a list of the top 50 most-cited articles from 2002 to 2022. Variables analyzed included citation count, annual citations, publication year, journal with impact factor, author details, country, institution, and funding.

**Results:** Citations for 50 articles varied from 67 to 919, with a median of 129 and an IQR of 210. Most of the articles (n = 34) were published between 2013 and 2018. The number of authors per article ranged from 3 to 18. Munir J.G. coauthored the most significant number of articles (n = 7). The top countries of origin for the articles were the UK (n = 15), the USA (n = 10), and Australia (n = 7). The dietary interventions discussed in our list included low FODMAP (n = 27), probiotics (n = 6), GFD (n = 4), an Ig4 exclusion diet (n = 3), and other interventions (n = 10). Female authors were the first authors in 30 articles and senior authors in 19 articles, compared to their male counterparts.

**Conclusions:** An analysis of the top 50 cited articles on nutritional interventions in IBS highlights interest and impact trends, aiding policymakers in funding decisions. The predominance of a low-FODMAP diet underscores the focus on efficacy, potentially guiding standardized dietary guidelines.

## Introduction

Irritable Bowel Syndrome (IBS) is a gastrointestinal disorder that affects approximately 4% of the world’s population, with a two-fold higher prevalence among women [1, 2]. Given that the pathophysiology of the disease is multifactorial and unclear, it is well documented that patients with IBS have limited treatment options and exhibit poor quality of life linked to more frequent use of healthcare services as an additional financial burden [3].

Individuals with IBS often link their symptoms to particular foods, which results in diet restriction that may lead to nutritional deficiencies and malnutrition [4, 5]. Nutritional interventions, especially the dietary restriction of fermentable carbohydrates (commonly termed the ’low FODMAP (fermentable oligosaccharides, disaccharides, monosaccharides, and polyols) diet’), have gained popularity in treating IBS [6–8]. Dietary components significantly impact the gut microbiome composition and can subsequently affect IBS symptoms. Exploring nutritional interventions can also offer valuable insights into how to use this interplay for the treatment of IBS. There have been several publications on the most-cited articles related to IBS [9–14]; however, none of these listings specifically focused on nutritional interventions for IBS, and there is a lack of evidence-based dietary guidelines for the management of IBS. Therefore, our research can help establish standardized nutritional recommendations for patients.

*Bibliometric analysis* is an easy-to-understand analytical method used to assess the trends and frequency of citations in the published literature.

Evidence-based medicine requires practicing physicians to be up-to-date with relevant research in their field to best serve patients, according to the latest guidelines. Bibliometrics provide physicians with a body of knowledge and a collection of the most influential research papers in their field [15]. Citation analysis can help direct medical research funding to specific areas needing increased research activity by examining research articles’ relevance, significance, and impact [16]. To fill the existing gap in knowledge, we have carried out a targeted bibliometric analysis to assess the most significant articles about this area of study.

## METHODS

### Literature Search and Search Strategy

We used the Scopus database to retrieve the relevant articles for our bibliometric analysis. Published analyses and comparisons have demonstrated that, in the context of scientific publications, Scopus exhibits a broader scope of coverage than alternatives such as PubMed, Web of Science, and Google Scholar [15, 17]. Two reviewers (Sadaf Iftikhar and Saad Ahmed) independently searched Scopus in December 2023, with a time restriction from 2002 to 2022. Any reviewer discrepancies were resolved by consulting the third reviewer (Faraz Arshad).

The search strategy to obtain articles on IBS and nutritional interventions involved using ((Irritable AND Bowel AND Syndrome) OR (Colitis AND Mucous) OR (Colon AND Irritable)) AND ((Diet OR Disaccharides OR Monosaccharide OR Simple AND Sugars)). All individual search terms and words were searched in the article titles, abstracts, and keywords.

### Inclusion and Exclusion Criteria

The initial compilation of articles retrieved from Scopus consists of those identified under the journal articles category. Two reviewers (Sadaf Iftikhar and Saad Ahmed) separated the original articles from the review studies. Exclusively including original articles allows for more direct analysis of primary research findings, enhancing the study’s credibility by minimizing potential biases introduced by interpretations or summaries found in review articles. Studies on non-human subjects, those without abstract availability, and those in languages other than English were excluded. The selected two-decade period ensures that our study incorporates the latest advancements in research on nutritional interventions for managing irritable bowel syndrome. Additionally, including all journals provides comprehensive coverage of the relevant literature, minimizing the risk of overlooking valuable research from diverse sources.

### Data extraction

The search yielded 29637 studies in total. The abstracts were then analyzed to assess their relevance to the topic and determine whether they were suitable for inclusion. The articles obtained were consolidated and systematically organized using the ’cited by’ option. The final list comprised the 50 most-cited articles. Any reviewer discrepancies were resolved by consulting the third reviewer (Faraz Arshad). The concordance rate between the reviewers was 95%. Citation analysis of the 50 most cited articles was conducted using Scopus, and the chosen articles were manually screened. Data on each publication were extracted, encompassing the total citation count, mean citations per year, trends of articles and citations over the years, publication year, journal name along with its impact factor (IF), author count, author’s H-index, gender of first and last author titles, references, keywords, country of origin, institutional affiliation, and funding information. The mean, median, and IQR were computed for the total number of citations and the number of citations per year. Journal impact factors were acquired from Thomson Reuters’ Journal Citation Reports. The gender of the first and senior authors was ascertained by reviewing their photographs on institutional websites or noting the pronouns used to address them. It is agreed upon in existing literature that the last authorship position is held by the author with the most significant degree of seniority; for that matter, we reviewed authors in the first and previous positions. When an article had a solitary author, that author was considered the senior author [18].

### Data Analysis

Tables and charts were generated using Microsoft Excel 2016. IBM SPSS Statistics 23.0 was employed to conduct the Spearman correlation test, which assessed the correlation between the journal impact factor and the total number of citations by papers from our 50 list within that journal. A p-value of <0.05 was considered statistically significant.

## Results

### Citation Count, Citation Per Year, Citation Trend

Table 1 shows the 50 most cited citations and total citations per year. The number of citations for these 50 articles ranged from 67 to 919, with a median of 129.00 and an interquartile range of 210. The citations for all articles total 10515. The number of citations per year ranged from 4.3 to 102. The median, mean number, and IQR of citations per year were 15.9, 23.3, and 20.2, respectively. Figure 1 shows the trend of the total citations of the 50 articles in the list by year. There was a rapid rise in the total number of citations, starting in 2004, which almost doubled in 2013, followed by a significant drop in 2015, after reaching a maximum in 2017. After a minor drop in 2018, a significant decrease occurred in 2019.

**Figure 1:**
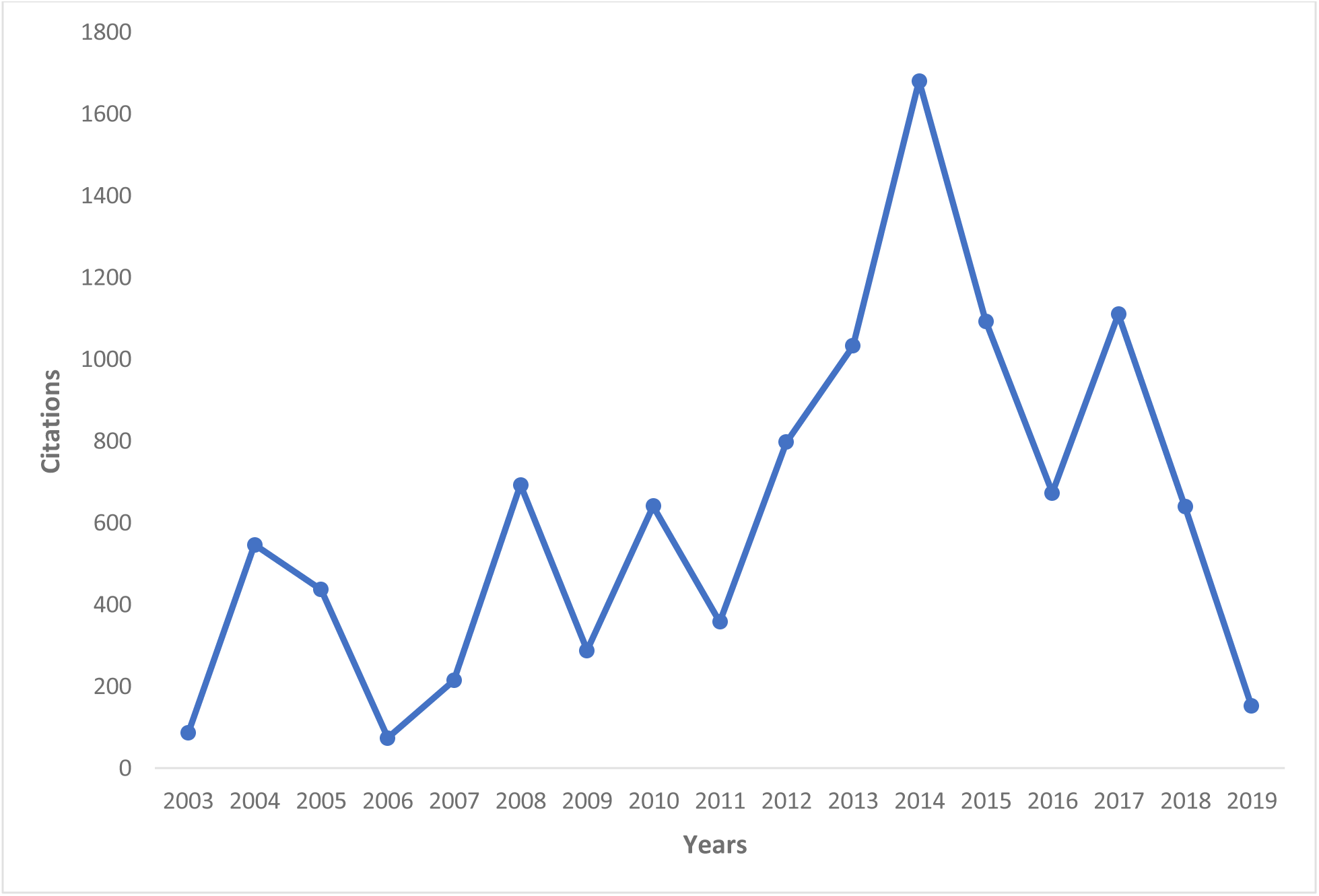
Total Citations of articles in the list each year.

**Table 1:** Top 50 original articles, their citations, citations per year, type of articles, and outcome.

| Rank | Article | Total citations | Avg citations per year | Type | Outcomes |
| --- | --- | --- | --- | --- | --- |
| 1 | A diet low in FODMAPs reduces symptoms of irritable bowel syndrome; 2014 | 919 | 102.1 | RCT | A low FODMAP diet is effective |
| 2 | Diets that differ in their FODMAP content alter the colonic luminal microenvironment; 2015 | 518 | 64.8 | RCT | FODMAP restriction to the level of symptom control should be excised |
| 3 | Manipulation of dietary short-chain carbohydrates alters the pattern of gas production and the genesis of symptoms in irritable bowel syndrome; 2010 | 490 | 37.7 | RCT | A low FODMAP diet is effective |
| 4 | Fermentable carbohydrate restriction reduces luminal bifidobacteria and gastrointestinal symptoms in patients with irritable bowel syndrome; 2012 | 462 | 42 | RCT | A low FODMAP diet is effective |
| 5 | Diet Low in FODMAPs Reduces Symptoms of Irritable Bowel Syndrome as Well as Traditional Dietary Advice: A Randomized Controlled Trial; 2015 | 458 | 57.3 | RCT | A combination of diet low in FODMAP with a traditional diet is effective |
| 6 | Food elimination based on IgG antibodies in irritable bowel syndrome: A randomized controlled trial; 2004 | 452 | 23.8 | RCT | Exclusion of diet based on IgG antibodies is effective |
| 7 | Dietary Triggers of Abdominal Symptoms in Patients With Irritable Bowel Syndrome: Randomized Placebo-Controlled Evidence; 2008 | 451 | 30.1 | RCT | A low FODMAP diet is effective |
| 8 | Self-reported food-related gastrointestinal symptoms in IBS are common and associated with more severe symptoms and reduced quality of life; 2013 | 443 | 44.3 | Cross-sectional | A low FODMAP diet is effective |
| 9 | A controlled trial of a gluten-free diet in patients with irritable bowel syndrome-diarrhea: Effects on bowel frequency and intestinal function; 2013 | 382 | 38.2 | RCT | Gluten-free diet is beneficial |
| 10 | Comparison of symptom response following advice for a diet low in fermentable carbohydrates (FODMAPs) versus standard dietary advice in patients with irritable bowel syndrome; 2011 | 358 | 29.8 | Prospective cohort | A low FODMAP diet is effective |
| 11 | A randomized controlled trial of a probiotic combination VSL# 3 and placebo in irritable bowel syndrome with bloating; 2005 | 324 | 18 | RCT | Probiotic reduce abdominal bloating |
| 12 | FODMAPs alter symptoms and the metabolome of patients with IBS: A randomised controlled trial; 2017 | 313 | 52.2 | RCT | Low FODMAP diet is effective |
| 13 | A Diet Low in FODMAPs Reduces Symptoms in Patients With Irritable Bowel Syndrome and A Probiotic Restores Bifidobacterium Species: A Randomized Controlled Trial; 2017 | 296 | 49.3 | RCT | Low FODMAP diet is effective |
| 14 | Differential effects of FODMAPs (Fermentable Oligo-, Di-, Mono-Saccharides and Polyols) on small and large intestinal contents in healthy subjects shown by MRI; 2014 | 291 | 32.3 | RCT | Low FODMAP diet is effective exclude inulin and fructose |
| 15 | A Randomized Controlled Trial Comparing the Low FODMAP Diet vs. Modified NICE Guidelines in US Adults with IBS-D; 2016 | 271 | 38.7 | RCT | A low FODMAP diet is effective |
| 16 | Confocal endomicroscopy shows food-associated changes in the intestinal mucosa of patients with irritable bowel syndrome; 2014 | 242 | 26.9 | RCT | A low FODMAP diet is effective |
| 17 | {A figure is presented} Predictors of Clinical Response to Gluten-Free Diet in Patients Diagnosed With Diarrhea-Predominant Irritable Bowel Syndrome; 2007 | 214 | 13.4 | Observational cohort | A gluten-free diet is effective for IBS |
| 18 | Clinical trial: Lactobacillus plantarum 299v (DSM 9843) improves symptoms of irritable bowel syndrome; 2012 | 210 | 19.1 | RCT | Probiotics are effective in treating IBS Symptoms |
| 19 | Soluble or insoluble fiber in irritable bowel syndrome in primary care? Randomized placebo-controlled trial; 2009 | 186 | 13.3 | RCT | Psyllium is effective in IBS |
| 20 | Effects of varying dietary content of fermentable short-chain carbohydrates on symptoms, fecal microenvironment, and cytokine profiles in patients with irritable bowel syndrome; 2017 | 163 | 27.2 | RCT | A low FODMAP diet is effective |
| 21 | Multivariate modeling of fecal bacterial profiles of patients with IBS predicts responsiveness to a diet low in FODMAPs; 2018 | 161 | 32.2 | RCT | A low FODMAP diet is effective |
| 22 | Many Patients With Irritable Bowel Syndrome Have Atypical Food Allergies Not Associated With Immunoglobulin E; 2019 | 152 | 38 | Prospective cohort | Food allergen exclusion is effective |
| 23 | Long-term impact of the low-FODMAP diet on gastrointestinal symptoms, dietary intake, patient acceptability, and healthcare utilization in irritable bowel syndrome; 2018 | 146 | 29.2 | RCT | A low FODMAP diet is effective |
| 24 | eHealth: Low FODMAP diet vs Lactobacillus rhamnosus GG in irritable bowel syndrome; 2014 | 138 | 15.3 | RCT | Both FODMAP and probiotics is effective |
| 25 | A double-blind randomized controlled trial of a probiotic combination in 100 patients with irritable bowel syndrome; 2008 | 130 | 8.7 | RCT | Probiotics is not superior to placebo |
| 26 | Prevalence and Presentation of Lactose Intolerance and Effects on Dairy Product Intake in Healthy Subjects and Patients With Irritable Bowel Syndrome; 2013 | 128 | 12.8 | RCT | Lactose-containing food should be eliminated |
| 27 | Diet and effects of diet management on quality of life and symptoms in patients with irritable bowel syndrome; 2012 | 125 | 11.4 | Cohort study | A low FODMAP diet improves the quality of life |
| 28 | Non-celiac gluten sensitivity has narrowed the spectrum of irritable bowel syndrome: A double-blind randomized placebo-controlled trial; 2015 | 117 | 14.6 | RCT | A gluten-free diet is effective |
| 29 | Food-specific serum IgG4 and IgE titers to common food antigens in irritable bowel syndrome; 2005 | 113 | 6.3 | Cross-sectional | IgG food items are responsible for IBS SYMPTOMS and such food items are excluded |
| 30 | Fructose intolerance in IBS and utility of fructose-restricted diet; 2008 | 112 | 7.5 | Retrospective cohort | Fructose-restricted diet effective in IBS |
| 31 | Randomized clinical trial: the efficacy of gut-directed hypnotherapy is similar to that of the low FODMAP diet for the treatment of irritable bowel syndrome; 2016 | 110 | 15.7 | RCT | Gut-directed hypnotherapy is as effective as FODMAP therapy |
| 32 | Follow-up of patients with functional bowel symptoms treated with a low FODMAP diet; 2016 | 104 | 14.9 | Retrospective cohort | A low FODMAP diet is effective |
| 33 | A Very Low-Carbohydrate Diet Improves Symptoms and Quality of Life in Diarrhea-Predominant Irritable Bowel Syndrome; 2009 | 102 | 7.3 | RCT | A low FODMAP diet is effective |
| 34 | Association Between Ultra-Processed Food Consumption and Functional Gastrointestinal Disorders: Results From the French NutriNet-Santé Cohort; 2018 | 99 | 19.8 | Cross-sectional | Ultra-processed food consumption is associated with IBS so UPF should be excluded |
| 35 | Efficacy of a Gluten-Free Diet in Subjects With Irritable Bowel Syndrome-Diarrhea Unaware of Their HLA-DQ2/8 Genotype; 2016 | 99 | 14.1 | RCT | A gluten-free diet is effective |
| 36 | Long-term irritable bowel syndrome symptom control with reintroduction of selected FODMAPs; 2017 | 97 | 16.2 | RCT | A low FODMAP diet is effective |
| 37 | Self-management for women with irritable bowel syndrome; 2004 | 94 | 4.9 | RCT | Multicomponent intervention improves quality of life in IBS |
| 38 | Dietary sorbitol and mannitol: Food content and distinct absorption patterns between healthy individuals and patients with irritable bowel syndrome; 2014 | 91 | 10.1 | RCT | Dietary restriction of polyols is effective in IBS |
| 39 | Effects of scFOS on the composition of fecal microbiota and anxiety in patients with irritable bowel syndrome: a randomized, double-blind, placebo-controlled study; 2017 | 89 | 14.8 | RCT | Short-chain fructooligosaccharide is effective in IBS |
| 40 | Randomized clinical trial: low-FODMAP rye bread vs. regular rye bread to relieve the symptoms of irritable bowel syndrome; 2016 | 89 | 12.7 | RCT | A low FODMAP diet is effective in IBS |
| 41 | Irritable bowel syndrome in primary care: The patient's and doctor's views on symptoms, etiology and management; 2003 | 86 | 4.3 | Cross-sectional | Patients and GP have different perspectives of dietary intervention |
| 42 | Volatile Organic Compounds in Feces Associate With Response to Dietary Intervention in Patients With Irritable Bowel Syndrome; 2018 | 84 | 16.8 | RCT | A low FODMAP diet is effective |
| 43 | Effects of Prebiotics vs a Diet Low in FODMAPs in Patients With Functional Gut Disorders; 2018 | 82 | 16.4 | RCT | Prebiotics as effective as the low FODMAP diet |
| 44 | A candidate probiotic with unfavorable effects in subjects with irritable bowel syndrome: A randomized controlled trial; 2010 | 81 | 6.2 | RCT | Probiotics are unfavorable in IBS symptoms |
| 45 | IgG-based elimination diet in migraine plus irritable bowel syndrome; 2013 | 80 | 8 | RCT | IgG-based dietary elimination is effective |
| 46 | Randomized clinical trial: the analgesic properties of dietary supplementation with palmitoylethanolamide and polydatin in irritable bowel syndrome; 2017 | 78 | 13 | RCT | The dietary supplement palmitoylethanolamide/ polydatin for abdominal pain in patients with IBS suggests that this is a promising natural supplement. |
| 47 | A Diet Low in Fermentable Oligo-, Di-, and Monosaccharides and Polyols Improves Quality of Life and Reduces Activity Impairment in Patients With Irritable Bowel Syndrome and Diarrhea; 2017 | 74 | 12.3 | RCT | A diet low in FODMAPs led to significantly greater improvements in health-related QOL, anxiety, and activity impairment compared with a diet based on traditional recommendations for patients with IBS-D |
| 48 | Treating Irritable Bowel Syndrome with a Food Elimination Diet Followed by Food Challenge and Probiotics; 2006 | 74 | 4.4 | Open-label pilot study | The FODMAP diet is more effective than the traditional diet recommended by NICE |
| 49 | Kiwifruit improves bowel function in patients with irritable bowel syndrome with constipation; 2010 | 70 | 5.4 | RCT | Kiwi fruit consumption is beneficial for bowel function in IBS |
| 50 | Low fermentable oligo-di-mono-saccharides and polyols diet versus general dietary advice in patients with diarrhea-predominant irritable bowel syndrome: A randomized controlled trial; 2018 | 67 | 13.4 | RCT | Low FODMAP diet and general dietary advice both are beneficial for IBS and low FODMAP is more beneficial for IBS |

### Journal, institutional affiliation, and Funding Sponsor

The 50 most-cited articles were published in nine different journals. The top four journals produced over half of the articles. The top 4 journals include Clinical Gastroenterology and Hepatology (n = 8, 16%), followed by Gastroenterology (n = 7, 14%), American Journal of Gastroenterology (n = 5, 10%), and Gut (n = 4, 8%). Table 2 shows journals that have included more than one article in the most cited 50.

**Table 2:** Top Journals with Impact Factor and number of articles.

| Journal's name | Number of articles | 2022 Impact Factor |
| --- | --- | --- |
| Clinical Gastroenterology And Hepatology | 8 | 12.6 |
| Gastroenterology | 7 | 29.4 |
| American Journal Of Gastroenterology | 5 | 12 |
| Gut | 4 | 24.5 |
| Neurogastroenterology And Motility | 4 | 3.5 |
| World Journal Of Gastroenterology | 4 | 4.3 |
| Alimentary Pharmacology And Therapeutics | 3 | 7.6 |
| Journal Of Gastroenterology And Hepatology Australia | 2 | 4.1 |
| Journal Of Human Nutrition And Dietetics | 2 | 3.3 |

The IF of these journals ranged from 3.3 to 29.4. We found a direct and significant association between the journal impact factor and several citations received by articles from our list published in that journal with a p-value of 0.04 using the Spearman correlation test. (Figure 2)

**Figure 2:**
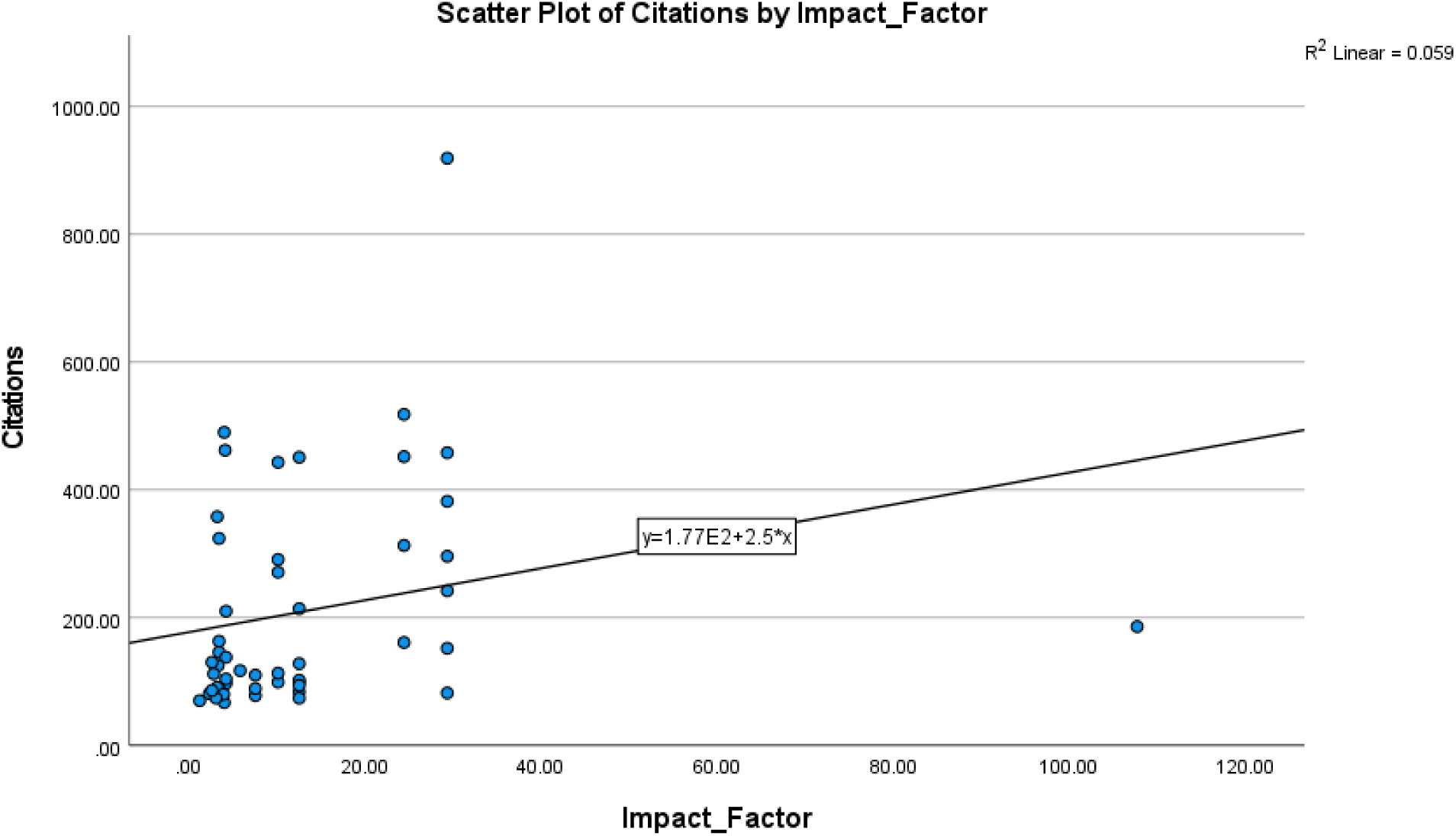
Scatter Plot of citations by Impact Factor.

Our list of articles featured a diverse range of affiliations with various institutions. Notably, Monash University led with seven articles. Following closely behind it were the Guys and St Thomas NHS Foundation, the Faculty of Medicine, Nursing, and Health Sciences, with five publications, and Hospital Universitario Vall Hebron, with four publications, as shown in Figure 3.

**Figure 3:**
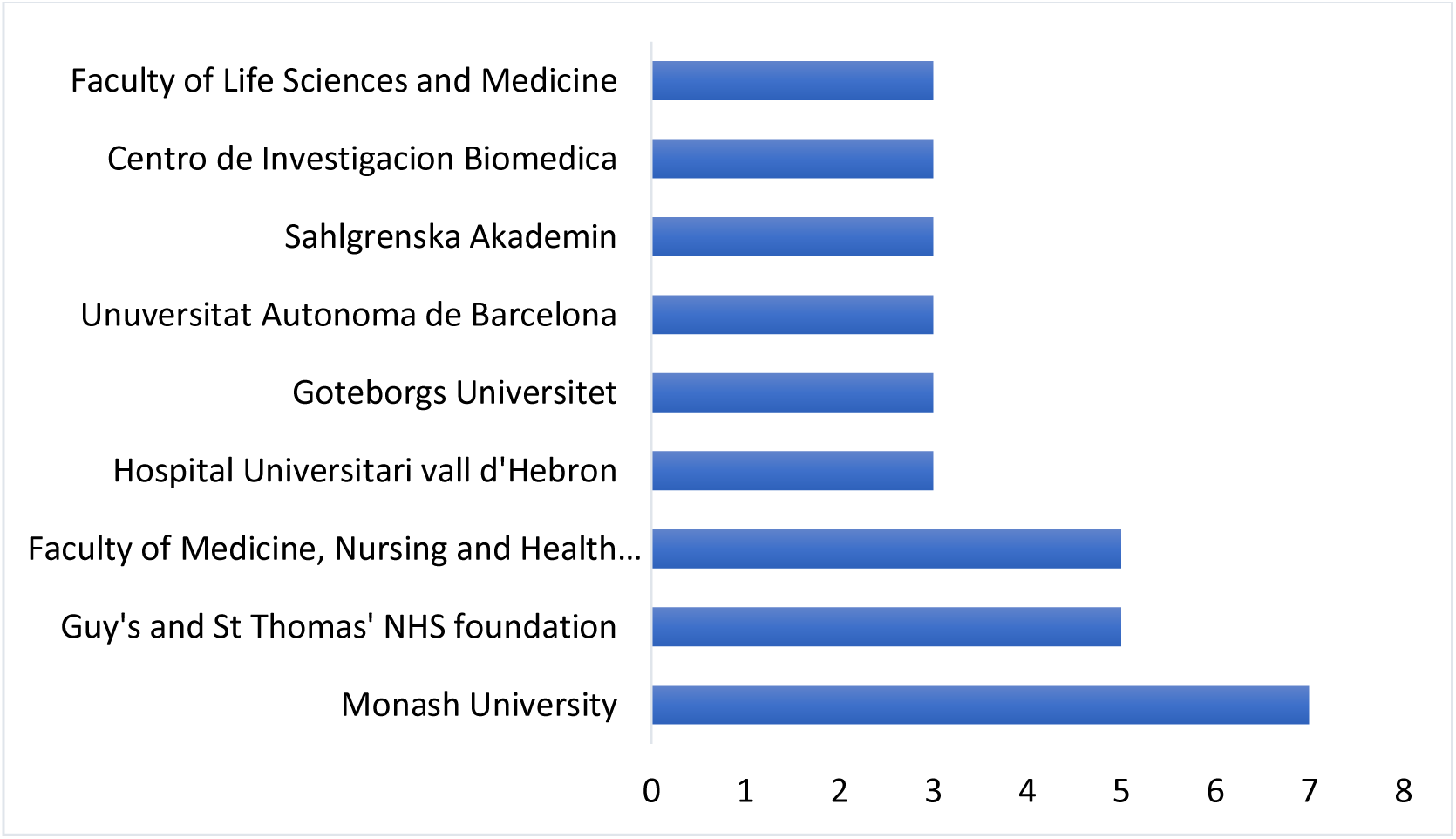
Institutions affiliated with articles.

The National Institute of Diabetes and Digestive Kidney Disease and the National Institute of Health have contributed to the funding of three articles and are closely followed by the Faculty of Medicine, Nursing, and Health Sciences at Monash University, Goteborgs University, and the Marianne and Marcus Wallenberg Foundation with two articles, as shown in Figure 4.

**Figure 4:**
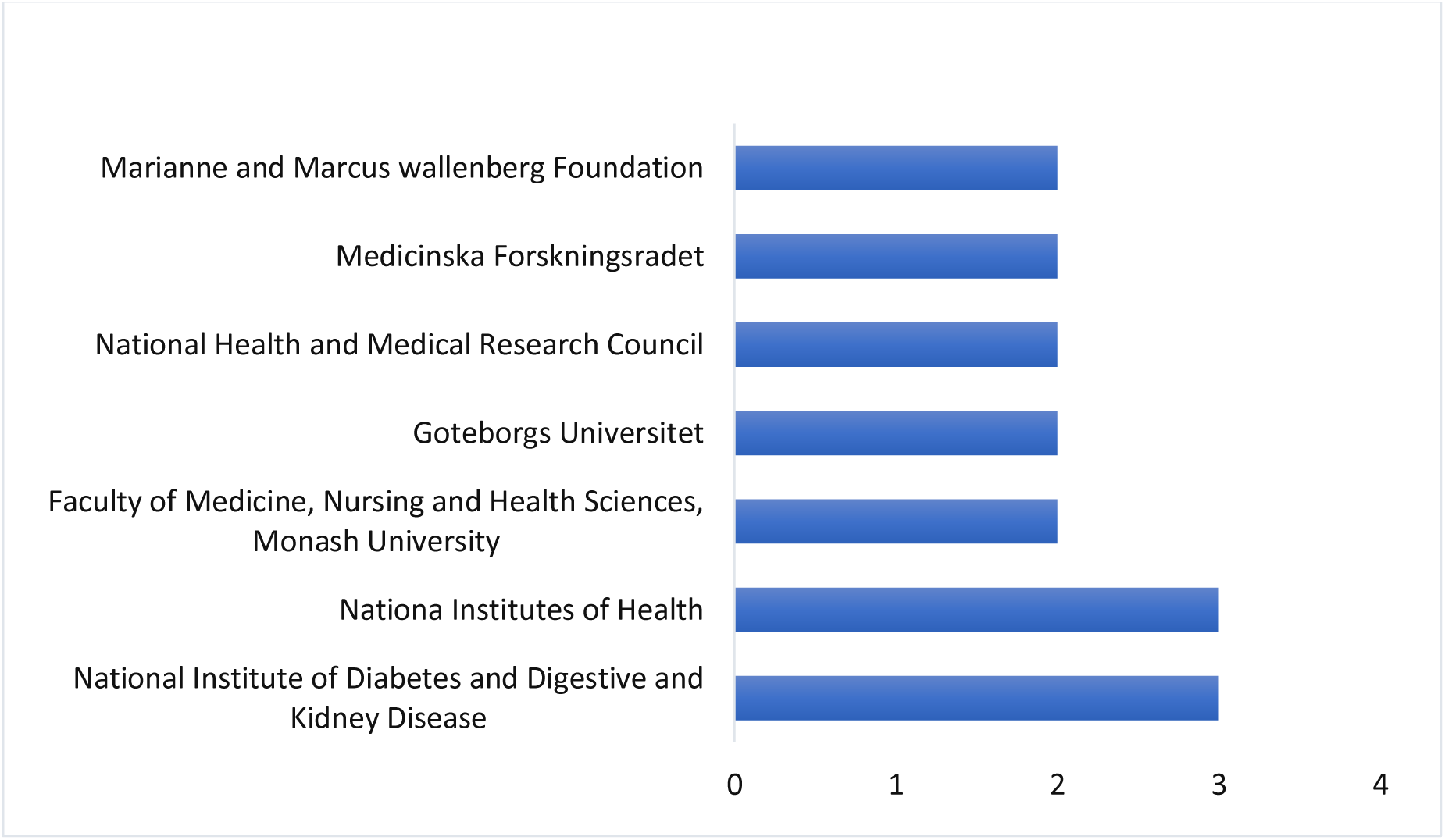
Sources of Funding.

### Year of Publication, Origins, and Authorship

The 50 most cited articles were published between 2002 and 2022. During these 20 years, the most significant number of articles (n = 34,68%) were published in five years, from 2013 to 2018 (Figure 5). The 50 most-cited papers originated from 8 countries, with over 15 articles from the United Kingdom. The three most popular countries of origin were the UK (n = 15,30%), the USA (n = 10,20%), and Australia (n = 7,14%) (Figure 6). A vast range of 263 authors contributed to 50 articles, and the number of authors per article ranged from 3 to 18. Each paper has a median of 7 authors with an interquartile range of 4. The authors with five or more articles are shown in Figure 7. The most significant number of most-cited articles were coauthored by Munir J.G., numbering (n = 7,14%) articles, followed by Gibson P.R. with (n = 6,12%) articles, Irving P.M., Staudacher H.M., and Whelan K, numbering (n = 5,10) articles, as shown in Table 3. The authors’ H-index highlights the role of the author in the scientific world as determined by the author-specific citation frequency, which is also included in Table 3. The H-index ranged from 26 to 82, among which Gibson, P.R., had the highest H index of 82.

**Figure 5:**
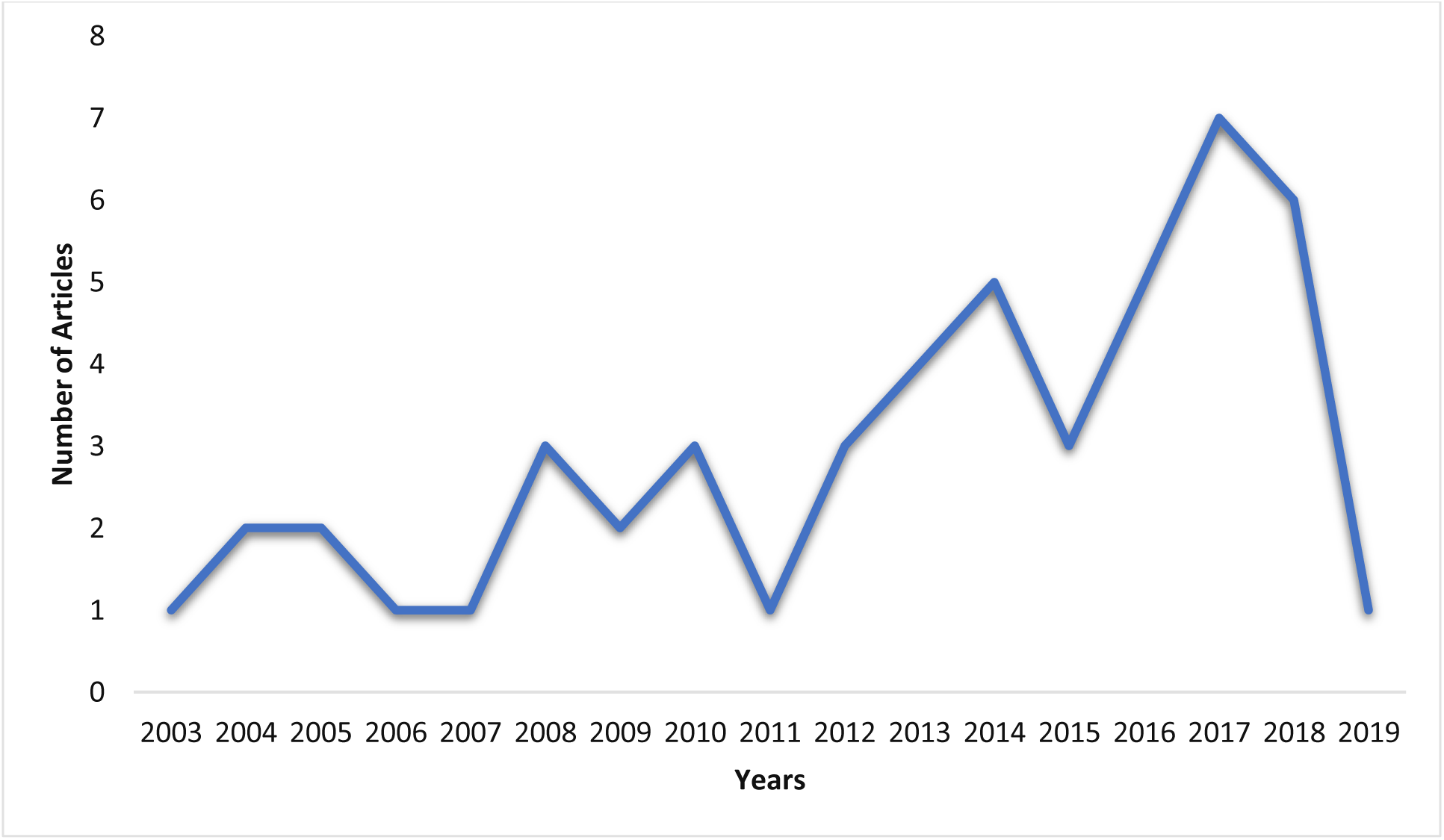
Number of articles in our list each year.

**Figure 6:**
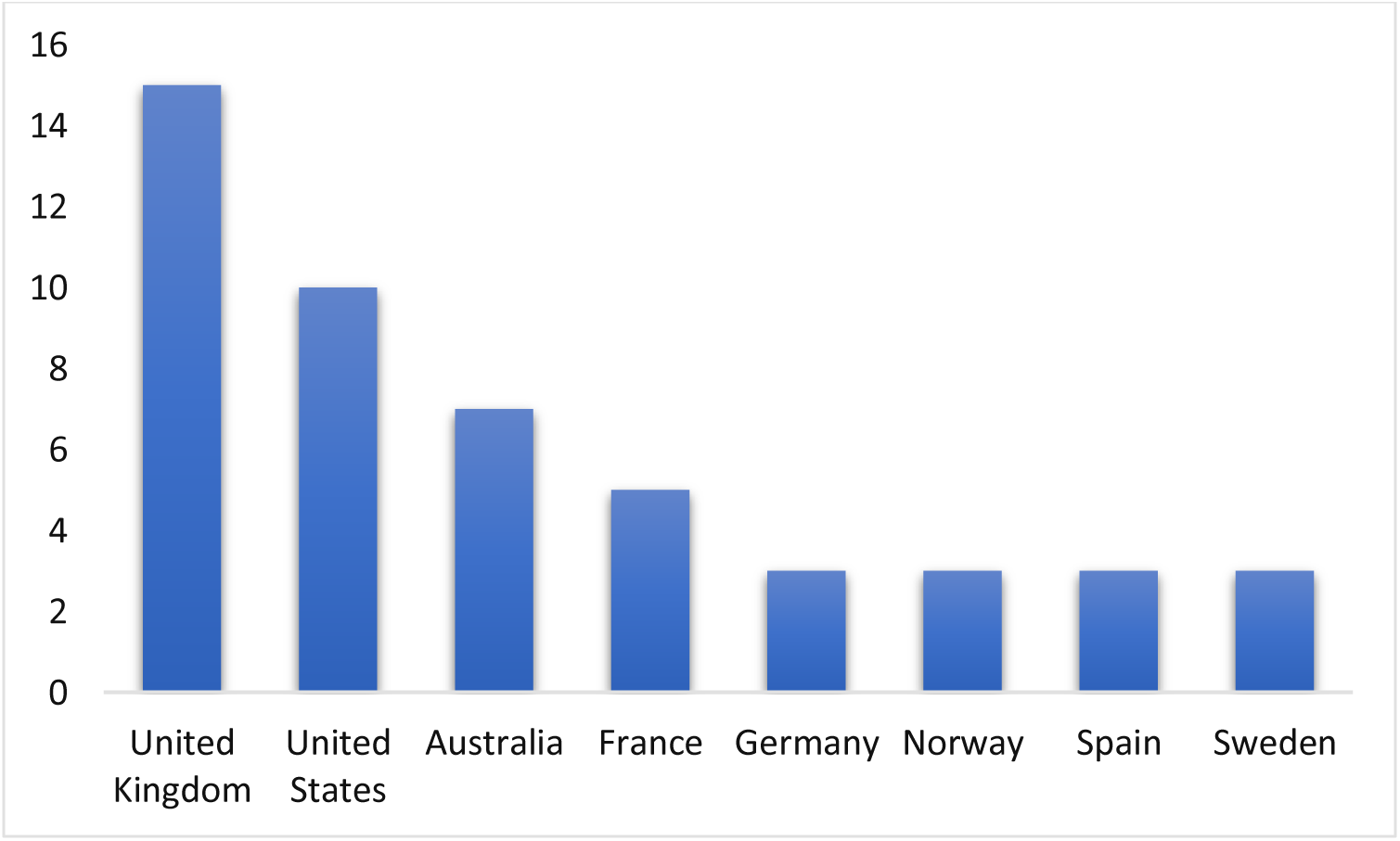
Number of articles originating from each country.

**Figure 7:**
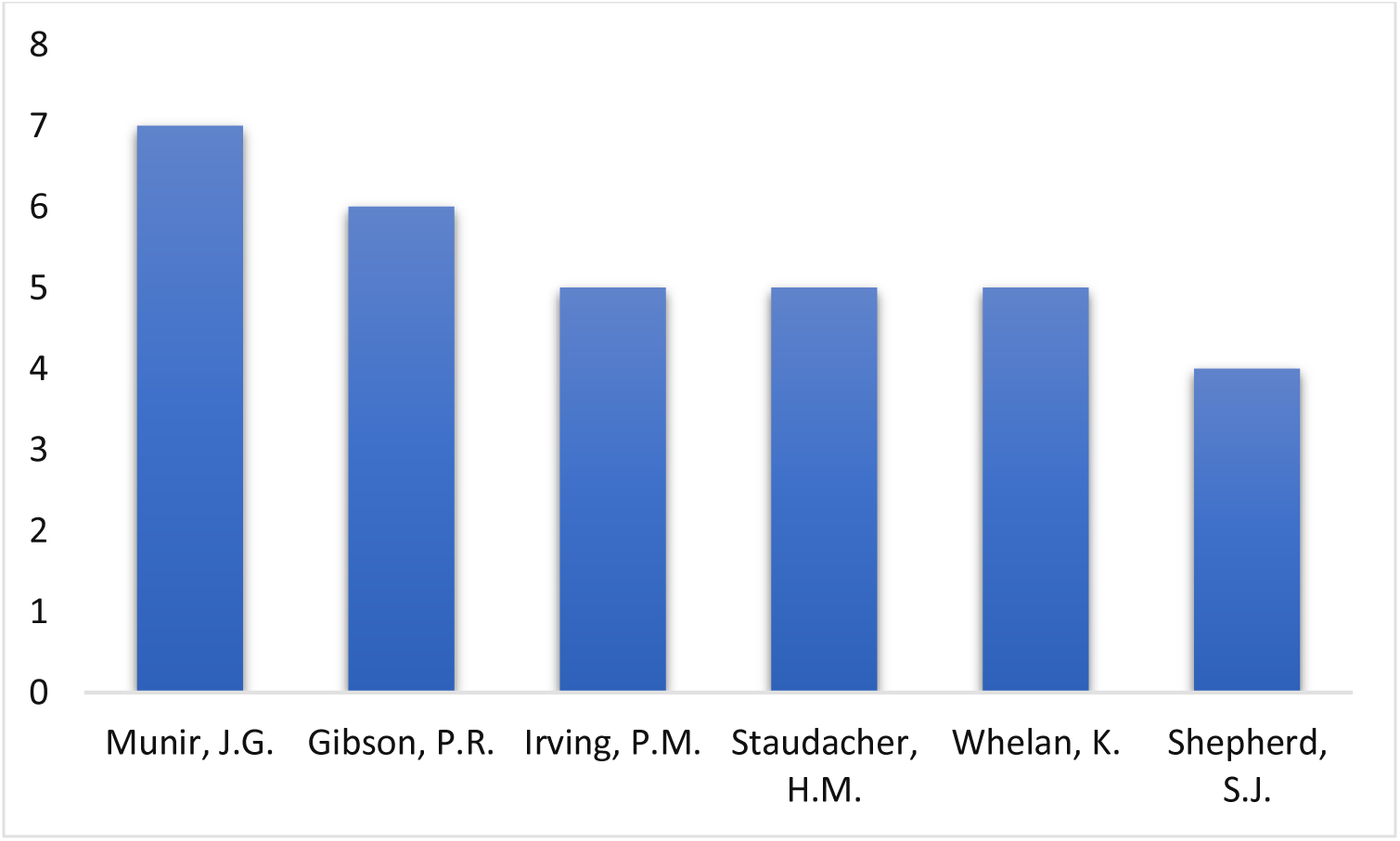
Number of articles by Top authors.

**Table 3:** Top authors with authorship position, Affiliation, Gender, and H-index.

| Author Name | Total articles | First position | Last position | Other position | Affiliation | Gender | H-index |
| --- | --- | --- | --- | --- | --- | --- | --- |
| Muir, J.G. | 7 | 0 | 4 | 3 | Monash University | F | 54 |
| Gibson, P.R. | 6 | 0 | 2 | 4 | Monash University | M | 82 |
| Irving, P.M. | 5 | 0 | 0 | 5 | Guy's and St Thomas' NHS Foundation Trust | M | 59 |
| Staudacher, H.M. | 5 | 3 | 0 | 2 | Deakin University | F | 26 |
| Whelan, K. | 5 | 0 | 3 | 2 | King's College London | M | 58 |
| Shepherd, S.J. | 4 | 1 | 0 | 3 | La Trobe University | F | 28 |

### Topical distribution

Nutritional interventions discussed in the 50 most cited articles are shown in the last column of Table 1. Among the 50 articles, a low FODMAP diet is discussed in (n = 27,54%), followed by probiotics mentioned in (n = 6,12%) articles, a gluten-free diet in (n = 4,8%) articles, an Ig4 exclusion diet in (n = 3,6%) articles, and other dietary approaches in (n = 10,20%) articles. (Figure 8) Other dietary options include:

- Psyllium.
- Lactose-restricted diet.
- Fructose-restricted diet.
- Ultra-processed food elimination.
- Polyols-restricted diet.
- Short-chain fructooligosaccharide.
- Palmitoylethanolamide/ppolydarin supplement.
- Kiwi fruit.
- Food allergen exclusion.

**Figure 8:**
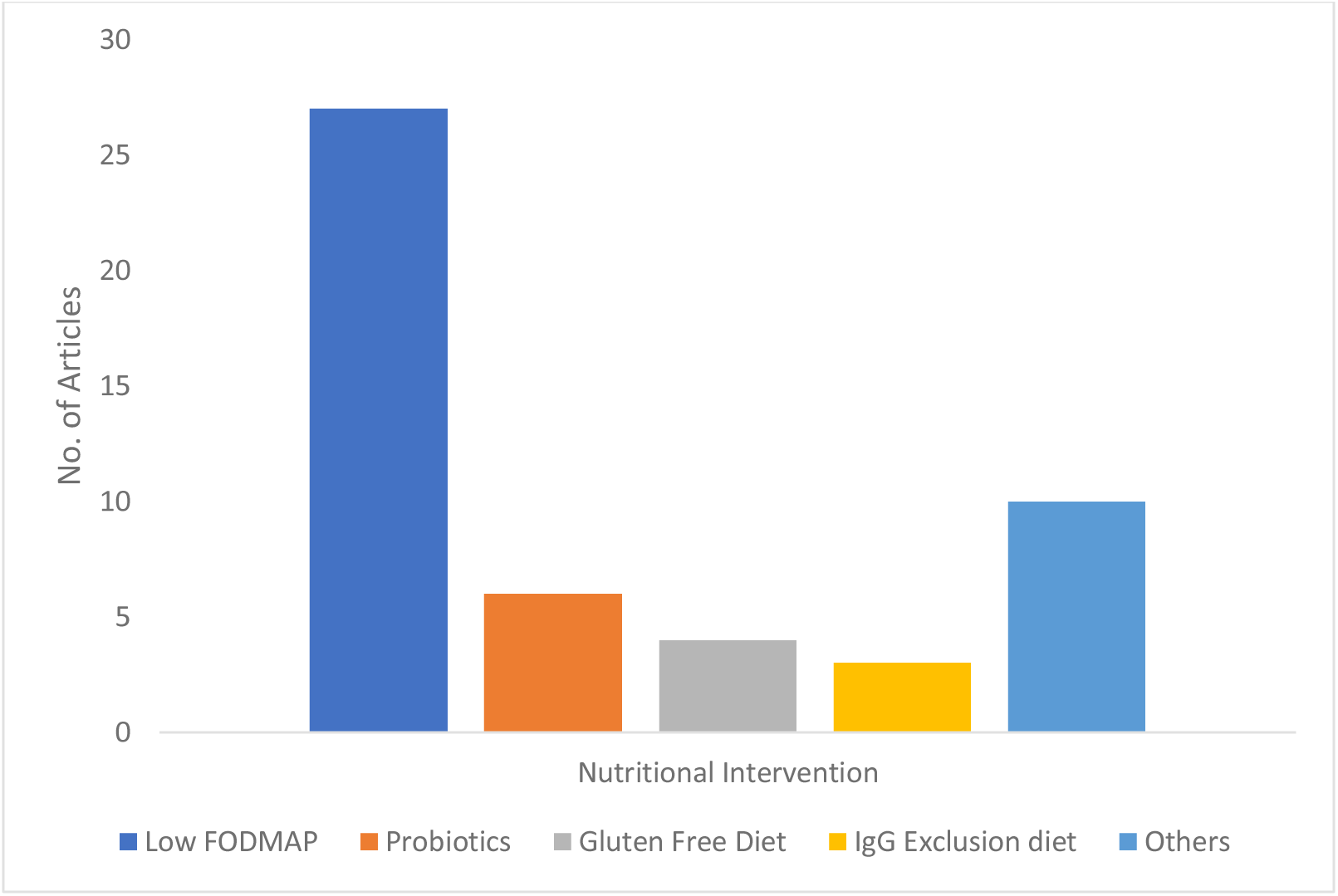
Nutritional Intervention discussed in articles.

### Gender of the senior and first authors

The gender of the first author and the senior author was determined in all 50 articles. Among first authors, females outnumbered males by a ratio of 3:2, with female authors (n = 30, 60%) and male authors (n = 20, 40%). On the other hand, it was the opposite for senior author distribution, with male authors (n = 31,62%) and female authors (n = 19,38%), as shown in Figure 9.

**Figure 09:**
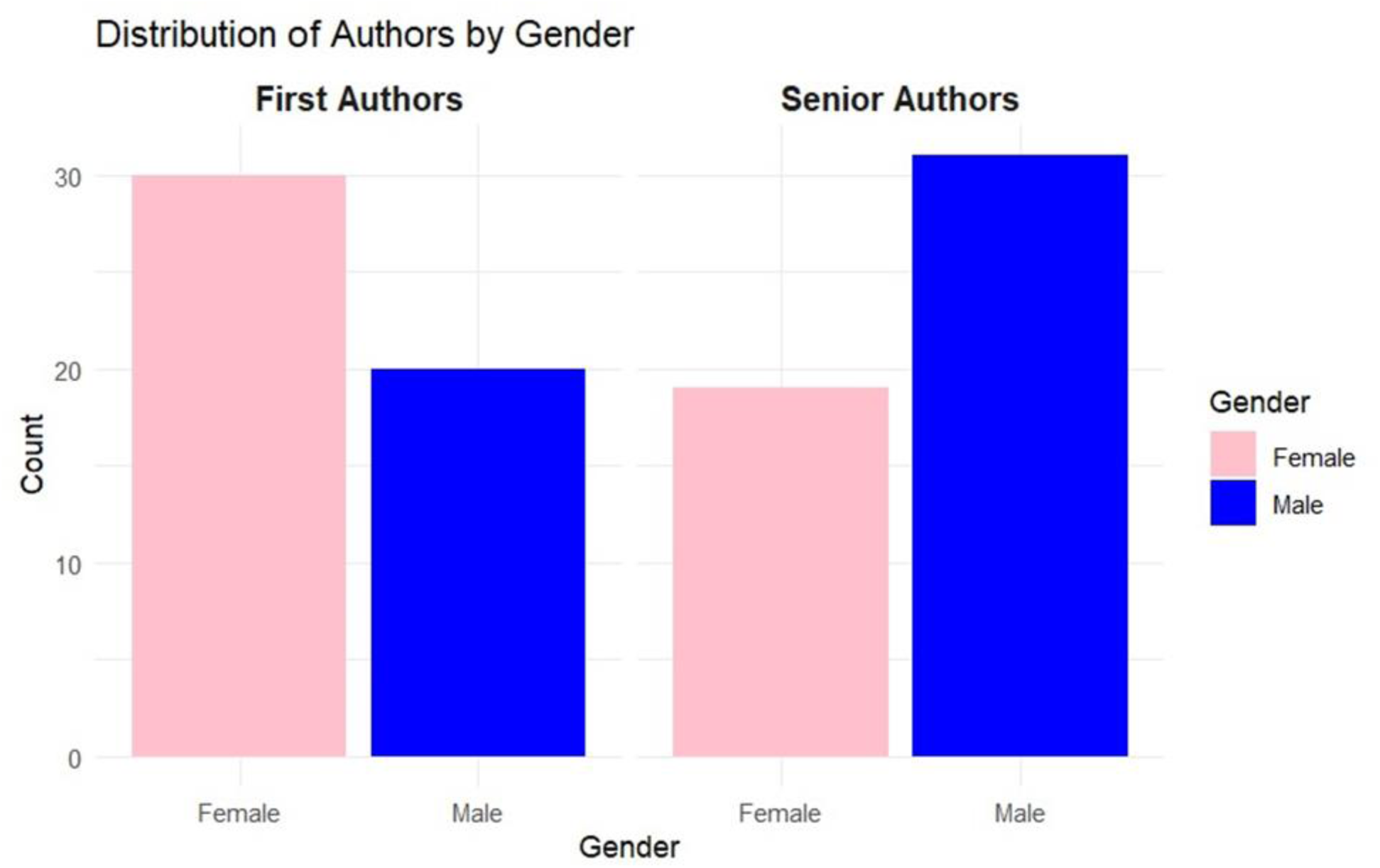
Distributions of authors by Gender.

## Discussion

IBS is a chronic functional disorder with recurrent abdominal pain associated with defecation and altered bowel habits [19]. Diagnosing IBS is challenging because it is typically a diagnosis of exclusion, which can prolong the diagnostic process and add to patient stress. As physicians, we must effectively educate our patients about this condition [20]. Understanding the reasons behind their symptoms is essential for patients with IBS, especially when no structural disease is present, as it greatly influences their management and well-being [21].

IBS presents as a heterogenous disorder, and efforts to develop targeted drug treatments have faced challenges due to uncertainties surrounding the nervous system in the gastrointestinal tract and central nervous system mechanisms contributing to visceral hypersensitivity in IBS. [22–24]. Combining behavioral strategies, dietary adjustments, and appropriate medication regimens maximizes successful treatment chances [25]. The sole purpose of this bibliometric analysis is to provide a comprehensive review of the literature surrounding nutritional interventions for IBS. Dietary therapy has recently gained popularity because research has highlighted that particular foods exacerbate or decrease the symptoms of IBS (26, 27).

The citation analysis depicted in Figure 1 indicates that the prime period of research engagement in nutritional interventions in IBS was from 2012–2017; during this timeframe, articles listed in the top 50 were the most frequently cited. This phase of rapid expansion can be analyzed to understand factors contributing to increased interest, such as breakthrough findings or emerging trends. This information can be instrumental for researchers, policymakers, and funding bodies to strategically plan and support ongoing efforts in nutritional intervention for IBS. The subsequent years witnessed fluctuations, with notable minimal citations in 2019. Fluctuations may reflect changes in public health priorities, guiding the development of policies that respond to current needs.

Table 1 includes journals consistently incorporating articles with the highest citation rates. These journals fall under the category of ’core journals’ or ’zone 1’ journals according to Bradford’s Law [28], which states that "zone 1 journals are those that are most frequently cited in the literature of the particular field and so are likely to be of value to researchers within that discipline". Similar to other bibliometric analyses [29, 30, 31], we also identified a pronounced and direct association between the journal’s impact factor and the number of articles with the highest citation rate published in that journal. This association emphasizes that a journal’s impact factor directly influences the number of citations its articles receive. Publishing in journals with high citation rates increases the visibility of research.

FODMAPs are carbohydrates poorly absorbed in the small intestine and can ferment in the large intestine, leading to symptoms such as bloating, gas, abdominal pain, and altered bowel habits. The mechanisms by which the FODMAP diet causes these symptoms include elevating osmotic pressure in the large intestine, acting as prebiotics for gas-producing Clostridium bacteria, serving as a substrate for bacterial fermentation leading to abdominal distension and discomfort, and interacting with enteroendocrine cells responsible for regulating GI sensation, motility, secretion, and absorption [32]. This leads to GI wall stretches and activates sensory wall receptors, leading to sensations of fullness, distension, and abdominal pain [33]. A low FODMAP diet improves the IBS symptom severity score (IBS-SSS) and the quality of life-related to the traditional diet [34]. Over half of the articles on our list focus on the FODMAP diet, highlighting its predominant importance in managing IBS. It’s noteworthy that maintaining a low-FODMAP diet requires detailed meal planning, involves significant expenses, and poses difficulties in adherence over an extended period (36, 37).

Probiotics, GFD, the Ig4 exclusion diet, and other dietary approaches are discussed less frequently, which indicates that these dietary options are an emerging area of research with growing interest but need to be established as low-FODMAP diets. Probiotics are safe and productive and improve gut health through their anti-inflammatory properties. They include Streptococcus thermophilus, the Lactobacillus strain, and Lactobacillus delbrueckii subsp. bulgaricus, and the Bifidobacterium strain [38]. They are efficacious and have gained popularity recently because they decrease bloating [39], decrease bowel movement per day [40], and lead to improvement on a 7-point Likert scale [41]. GFD includes gluten-free cereals like rice, buckwheat, corn, millet, quinoa, and various vegetables and fruits [42]. A recent trial suggests that FODMAP-restricted GFD improves quality of life and anxiety in IBS patients [43]. The elimination diet for IBS involves removing potential trigger foods from the diet and reintroducing them gradually to identify culprits. This process aims to customize dietary management for individuals with IBS, improving symptom control. (44) The elimination diet and other dietary options, while effective, are less reported in our 50-article list due to their complexity and the individualized approach required to identify specific food triggers for each patient.

Regarding national origin, we noted that the UK generated the most articles in the list of 50 most cited articles. Numerous European countries have pursued the lead set by the UK, with limited participation from Asian countries. Our results indicated the most significant number of most-cited articles were coauthored by Munir J.G., numbering seven, followed by Gibson P.R. with six articles. Considering academic and intellectual demands for recognizing individual impactful involvement, it may be advantageous to pinpoint and distinguish the most remarkable authors from front-runners. Scientists who make meaningful and evident contributions to the knowledge base experience heightened eminence and amplified influence. This influence extends to the publication realm, where editors are more inclined to accept their work and seek opinions on articles [45].

Most principal authors were male, highlighting the scarcity of female representation in senior author positions [46]. This statistic may be influenced by the age discrepancy between male and female colleagues in college faculty, leading to younger female colleagues still being in the initial stages of their professional journey. Our bibliometric analysis revealed a distinguished trend where female authors were more prevalent in the first author position than the senior author. This observation contrasts with findings from other bibliometric analyses (47, 48), which often report different patterns of gender distribution in authorship roles. Our study suggests that the gender of the senior author does not necessarily coincide with that of the first author. While there is a gradual increase in the number of female first and senior authors in gastroenterology, additional steps need to be taken at the grassroots level to tackle gender discrepancies and ensure better representation of female authors in academic literature [49].

## Limitations

Despite our sincere efforts to limit the bias, it is crucial to acknowledge the obstacles inherent in our bibliometric analysis. Firstly, we extracted articles from only one database, Scopus. It has been discovered that it tends to miss older citations [17, 50]. However, there is evidence that the Scopus database has a broader scope of coverage compared to alternatives such as PubMed, Web of Science, and Google Scholar. Additionally, Scopus’s extensive coverage across multidisciplinary fields, compared to PubMed, aligns with the scope of our research [51]. Secondly, issues about the self-citation effect on bibliometric analyses have been raised, especially while a small group of authors dominates high-frequency citations, as glaring in our eyes. Although research has addressed this problem, self-citations may only partially affect bibliometrics [52]. A potential source of bias is the time needed for newly influential articles to accumulate measurable citations. Thus, our list will not consider some currently published excessive-impact articles, despite their future importance [53].

Moreover, quotation counts and H-indices may vary from those received in other competing databases, introducing variability within the evaluation. Finally, the limitation of including only the top 50 articles was due to insufficient literature. This highlights the need for further research to broaden the scope and identify additional trends.

## Conclusions

Analyzing the 50 most-cited articles in Nutritional Intervention in Irritable Bowel Syndrome provides valuable insights. The study of citation counts and trends indicates the level of interest and impact of research in nutritional interventions for IBS over time. Policymakers can use this data to prioritize funding and support for dietary research and public health initiatives. The predominant focus on low-FODMAP diets among the most cited articles suggests a strong interest in understanding the efficacy of nutritional interventions in managing IBS symptoms. This finding could lead to the development of standardized dietary guidelines for IBS patients. The domain of nutritional intervention for IBS has witnessed fluctuations in attention and acknowledgment over time, marked by phases of rapid expansion, decline, and stability. Assessing these trends can help identify research gaps; understanding these patterns can help policymakers and funding agencies decide where to allocate resources most effectively.

## Data Availability

All data produced in the present study are available upon reasonable request to the authors

## Abbreviations

IBS: Irritable Bowel Syndrome
IF: Impact Factor
GIT: Gastrointestinal Tract
FODMAP: fermentable oligosaccharides, disaccharides, monosaccharides, and polyols.
IBS-SSS: Irritable Bowel Syndrome Symptom Severity Score
GFD: Gluten-Free Diet
IQR: Interquartile Range

## Acknowledgments

Not applicable.

## Ethics declarations

### Ethics, approval, and consent to participate

Not applicable.

### Consent for publication

Not applicable.

